# Eye Exercises for Myopia Prevention and Control: A Systemic Review and Meta-Analysis

**DOI:** 10.1101/2023.01.29.23284986

**Authors:** Zhicheng Lin, Feng Xiao, Weiye Cheng

## Abstract

**Background:** Myopia is increasing in prevalence and developing at a younger age, a trend exacerbated by the COVID-19 pandemic. To combat the epidemic of myopia, eye exercises have been promoted in recent national efforts in mainland China, continuing a compulsory national school policy for over 50 years. We aimed to evaluate the efficacy of eye exercises in preventing and controlling myopia.

**Methods:** In this systemic review and meta-analysis, we searched nine major Chinese and English databases from their inception to December 15, 2022. We included studies that compared the effects of eye-exercise interventions with controls (no eye exercises) on at least one myopia-related indicator. Studies could be either randomized or non-randomized controlled trials. Two coders independently screened records for eligibility; extracted study-level data (study information, sample sizes, interventions, and myopia indicators); and assessed the risk of bias (Cochrane Risk of Bias Tool 2.0) and study heterogeneity (*I*^*2*^). Using random-effect models and sensitivity analysis, we estimated the effects of eye exercises compared to control on changes in visual acuity, diopter, and curative effects (axial length was not reported). We used standardized mean differences (SMDs) to evaluate visual acuity and diopter outcomes, and risk ratios (RRs) to assess curative effects. This study is registered on the Open Science Framework (https://osf.io/dr5jk).

**Findings:** Of the 1765 records identified, 1754 were excluded: 423 were duplicates, 1223 did not have a control group, 16 did not have full-text, and 92 did not fulfill other inclusion criteria. In total, 11 studies were included in the meta-analysis, with 921 participants (399 in eye-exercise interventions and 522 in control groups). Nine studies had some concerns of bias in at least two domains, and two studies had a high risk of bias in two domains. Seven studies used visual acuity to measure myopia; visual acuity declined after eye-exercise interventions (SMD=–0·67, 95% CI –1·28 to –0·07, *Z*=2·17, p=0·03) and the effect was not better than control (SMD=–0·50, 95% CI –1·16 to 0·16, *Z*=1·49, p=0·14). Two studies used diopter to measure myopia; the effect of eye-exercise interventions did not differ from control (SMD=–1·74, 95% CI –6·27 to 2·79, *Z*=0·75, p=0·45). Seven studies reported curative effects; eye exercises had a higher curative effect than control (RR=0·40, 95% CI 0·23–0·71, *Z*=3·13, p<0·01).

**Interpretation:** Eye exercises are not effective in preventing or controlling the progression of myopia, as measured by changes in visual acuity and diopter. A small positive effect is observed in curative effects, but the studies have high heterogeneity and potential publication bias, with major weaknesses in design (inadequate measures, small sample sizes, biases, failure to consider side effects, and failure to include established effective interventions as control). There is little evidence to support the continued use of eye exercises to manage myopia in schoolchildren.

**Funding:** Guangdong Basic and Applied Basic Research Foundation (2019A1515110574) and Shenzhen Fundamental Research Program (JCYJ20210324134603010).

**Research in context:** *Evidence before this study:* Myopia is a growing global public health challenge and has reached epidemic proportions in East and Southeast Asia. Given the large population of schoolchildren in these regions and the societal burden and personal costs of myopia, myopia control has become a top public health priority, particularly in mainland China. Schoolchildren in mainland China have been required to perform eye exercises twice a day for over 50 years; this compulsory policy has also been emphasized in recent national efforts to combat the myopia epidemic. We searched PubMed for meta-analyses of controlled trials that assessed the efficacy of eye exercises against myopia onset or its progression, using search terms related to “myopia” and “eye exercises”, but did not retrieve any from database inception until January 23, 2023.

*Added value of this study:* This study is the first meta-analysis of controlled trials examining the efficacy of eye exercises in preventing and controlling myopia. By including trials published in Chinese and English from database inception to December 15, 2022, the meta-analysis found that visual acuity declined after eye-exercise interventions (SMD=–0·67, 95% CI –1·28 to –0·07, *Z*=2·17, p=0·03) and the effect was not better than control (SMD=–0·50, 95% CI –1·16 to 0·16, *Z*=1·49, p=0·14), with a similar pattern in diopter measures (SMD=–1·74, 95% CI –6·27 to 2·79, *Z*=0·75, p=0·45). Additionally, the curative effect of eye-exercise interventions was higher than control (RR=0·40, 95% CI 0·23–0·71, *Z*=3·13, p<0·01). The meta-analysis also highlighted five major weaknesses in extant studies: inadequate measures, small sample sizes, biases, failure to consider side effects, and failure to include established effective interventions as control.

*Implications of all the available evidence:* The findings of this study, along with previous observational evidence, suggest that there is little support for using eye exercises to prevent myopia or control its progression. These results challenge the continued use of eye exercises as a policy to control myopia in schoolchildren and emphasize the need for rigorous research to establish their efficacy.

## Introduction

Myopia is a rapidly growing public health challenge, affecting more than 2 billion people currently—a number projected to grow to 5 billion by 2050, about half of the population worldwide.^1^ In East and Southeast Asia, it has become an epidemic where more than 80% of young adults are myopic—a rapid rise from 20–30% in the mid-20^th^ century.^2,3^ As it becomes more prevalent, it is also developing at a younger age.^3^ Early onset of myopia is strongly associated with high myopia in adulthood^4^—more than 50% of those with myopia onset at 7 or 8 years of age develop high myopia (versus less than 5% of those with onset at 12 years or older).^5^ High myopia is a common cause of vision impairment and blindness, as it heightens the risk of cataract, glaucoma, retinal detachment, and myopic macular degeneration.^6^ In economic impact, myopic macular degeneration and uncorrected myopia—the leading cause of vision impairment—were estimated to be responsible for about US$250 billion in lost global productivity in 2015.^7^ Myopia thus presents an enormous challenge for health services, from screening and providing spectacles to managing eye diseases.^8^

Because of its rapidly growing prevalence and its societal burden and personal costs (eg, reduced quality of life), myopia control has become a top public health priority in countries such as China. A national survey of ∼2.5 million Chinese students reported 52·7% of them to be myopic by the end of 2020: 80·5% in high school students, 71·1% in middle school students, 35·6% in primary school students, and 14·3% in six-year-olds.^9^ A large proportion of myopic schoolchildren have no refractive correction,^10^ which undermines their school learning and health—for example, according to a recent city-wide study the ratio was about 60% in a southern municipal city, Shantou;^11^ the ratio was even higher in migrant children, estimated to be 85%.^12,13^ China recently set up specific, numeric goals for preventing and controlling myopia—goals that became part of evaluation metrics for all provincial and local governments (see appendix p 1 for the timeline of recent national efforts). The target was to reduce the prevalence of myopia by at least 0·5% annually from 2018–2023 (for provinces with high prevalence, at least 1% annually), such that by 2030, the prevalence would be reduced to <70% in high school students, <60% in middle school students, <38% in primary school students, and 3% in six-year-olds. To achieve these goals, a suite of implementation requirements was made at the levels of family, school, medical institute, student, and government agency.^14,15^ Prominent among these were compulsory eye exercises^16^ for schoolchildren, to be performed twice a day during school days—a policy that was dated to the 1960s, based on eye acupressure from traditional Chinese medicine (for an introduction of its history and rationale, see appendix p 2). This requirement was reaffirmed in response to the adverse impact of COVID-19 in a renewed concerted national plan issued in 2021.

This compulsory school policy has affected schoolchildren in mainland China for more than half a century. Yet, there has been no meta-analysis of controlled trials to evaluate the efficacy of eye exercises in myopia prevention or control. Long overdue, this important question acquires particular urgency in light of the recent nationwide policy goals and the negative impacts of the COVID-19 pandemic. Even though these goals are considered modest,^17^ progress has been stunted because of pandemic lockdown measures from 2020 to 2022.^18^ Home confinement has been associated with a substantial myopic shift, particularly in young children (aged 6–8 years);^19^ for example, from grade 2 to grade 3, myopia incidence almost doubled from late 2019 to late 2020 (20·8%, with lockdown) compared with the same period from 2018 to 2019 (13·3%, without lockdown).^20^ The cumulative effects over the past three years (2020– 2022) are likely to be even more pronounced, creating unprecedented challenges for controlling myopia. Thus, it is now more important and urgent than ever to adopt effective, evidence-based measures to combat myopia development. A challenge in evaluating the effect of eye exercises, however, is that some studies on this topic are published in Chinese that are not indexed in English databases. Here, we aimed to evaluate the overall efficacy of eye exercises in preventing myopia and slowing its progression, by conducting a meta-analysis of studies that compared eye-exercise interventions with controls that did not use eye exercises. We searched nine Chinese and English databases from their inception to December 15, 2022 in accordance with the Preferred Reporting Items for Systematic Reviews and Meta-Analyses guidelines.^21^

## Methods

### Search strategy and selection criteria

We assessed the effectiveness of eye exercises against myopia onset or progression (figure 1). We searched nine databases, both Chinese (i.e., CNKI) and English (i.e., Web of Science, Google Scholar, EBSCO, PubMed, Cochrane Library, Science Direct, Scopus, and Embase), spanning from database inception to December 15, 2022. The search terms were “eye exercise” AND “myopia” in Chinese for the Chinese database and (“myopia” OR “short sightedness” OR “nearsightedness”) AND (“ocular gymnastics” OR “eye exercises” OR “eye exercise”) for the English databases (see appendix p 3 for a full list of the search terms used).

**Figure 1:**
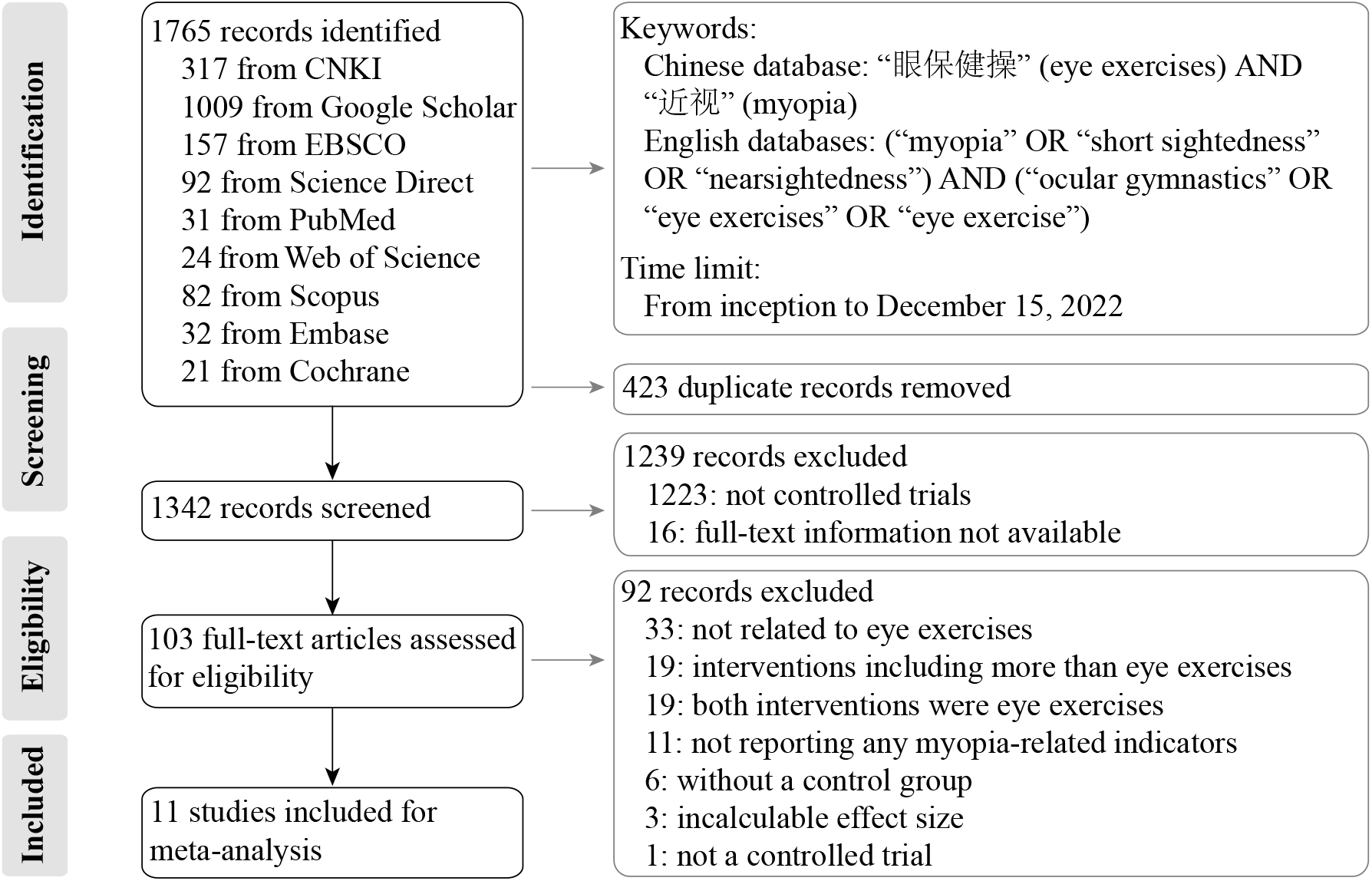
Study selection.

Four coders independently screened the literature: WC and YW from database inception to April 30, 2020; FX and XG from May 1, 2020 to December 15, 2022. The screening started with titles and abstracts first and then the full text, using the following selection criteria. Specifically, to be included, papers must meet all five criteria: (1) available as a journal publication or a dissertation; (2) using eye exercises for intervention; (3) employing a control group that did not use eye exercises; (4) reporting at least one myopia-related indicator (e.g., axial length, visual acuity, diopter, or curative effects); and (5) reporting data that enabled effect size extraction or estimation. Discrepancies between coders were resolved through discussion. All excluded articles during the full-text screening stage are listed in the appendix (pp 4–9). RevMan (version 5.4) was used to screen and organize articles.

### Data analysis

The analysis focused on outcome evaluation, risk of bias, and study heterogeneity. For each study, we extracted the article information, sample size, intervention(s), and myopia indicator(s). Of the four myopia indicators, visual acuity and diopter outcomes were evaluated using standardized mean differences (SMDs, with 95% CI), by dividing the mean difference between two groups with the standard deviation. Curative effects were assessed using the risk ratios (RRs, with 95% CI), by calculating the ratio of the risk of developing myopia (or progression) in the eye-exercise group to the risk in the control group. Axial length was not reported in any study.

Risk of bias was independently rated by two coders (FX and XG) using the Cochrane Risk of Bias Tool 2.0 (commonly recommended for randomized trials). Bias was rated across seven domains: random sequence generation; allocation concealment; blinding of participants and personnel; blinding of outcome assessment; incomplete outcome data; selective reporting; and other biases. Discrepancies in ratings were resolved after discussion. Intercoder consistency (reliability) was evaluated using linearly weighted PABAK (prevalence-adjusted, bias-adjusted kappa), which accounted for two characteristics of the ratings: 1) some ratings (e.g., some concerns) were much more prevalent than others (e.g., high risk), and 2) degree of disagreement differed among the three ratings (e.g., low and high risks were more different than low risk and some concerns).

Study heterogeneity was quantified using the *I*^*2*^ statistic (range from 0% to 100%, with higher values representing larger heterogeneity). The degree of heterogeneity was defined based on conventional standards: not substantial (*I*^*2*^<50%) or substantial (*I*^*2*^≥50%). When the heterogeneity was substantial, outsized influences of individual studies on the overall results were probed using sensitivity analysis (subgroup analysis was not appropriate given the small number of included papers). Random-effects models were used to summarize effect sizes. Analyses were conducted using RevMan (version 5.4) and R/RStudio (version 2022.07.1). Data and code are available online (https://osf.io/dr5jk/).

## Results

The initial search yielded 1765 articles, of which 1754 were excluded: 423 were duplicates, 1223 did not have a control group, 16 did not have full-text, and 92 did not fulfill other inclusion criteria (as detailed in figure 1). The list of excluded articles during the full-text screening stage is provided in the appendix (pp 4–9). In total, 11 studies (9 in Chinese and 2 in English) were included in the meta-analysis.

The 11 included studies^22-32^ were between 1974 and 2021; three were dissertations^23,24,29^. All were controlled trials, including 2 non-randomized controlled trials and 9 randomized controlled trials (table). They assessed three types of outcomes: visual acuity, diopter, and curative effect. In total, the meta-analysis included 921 participants: 399 in eye-exercise groups and 522 in control groups. All studies were conducted in children except one (6 to 26 years old).

**Table:**
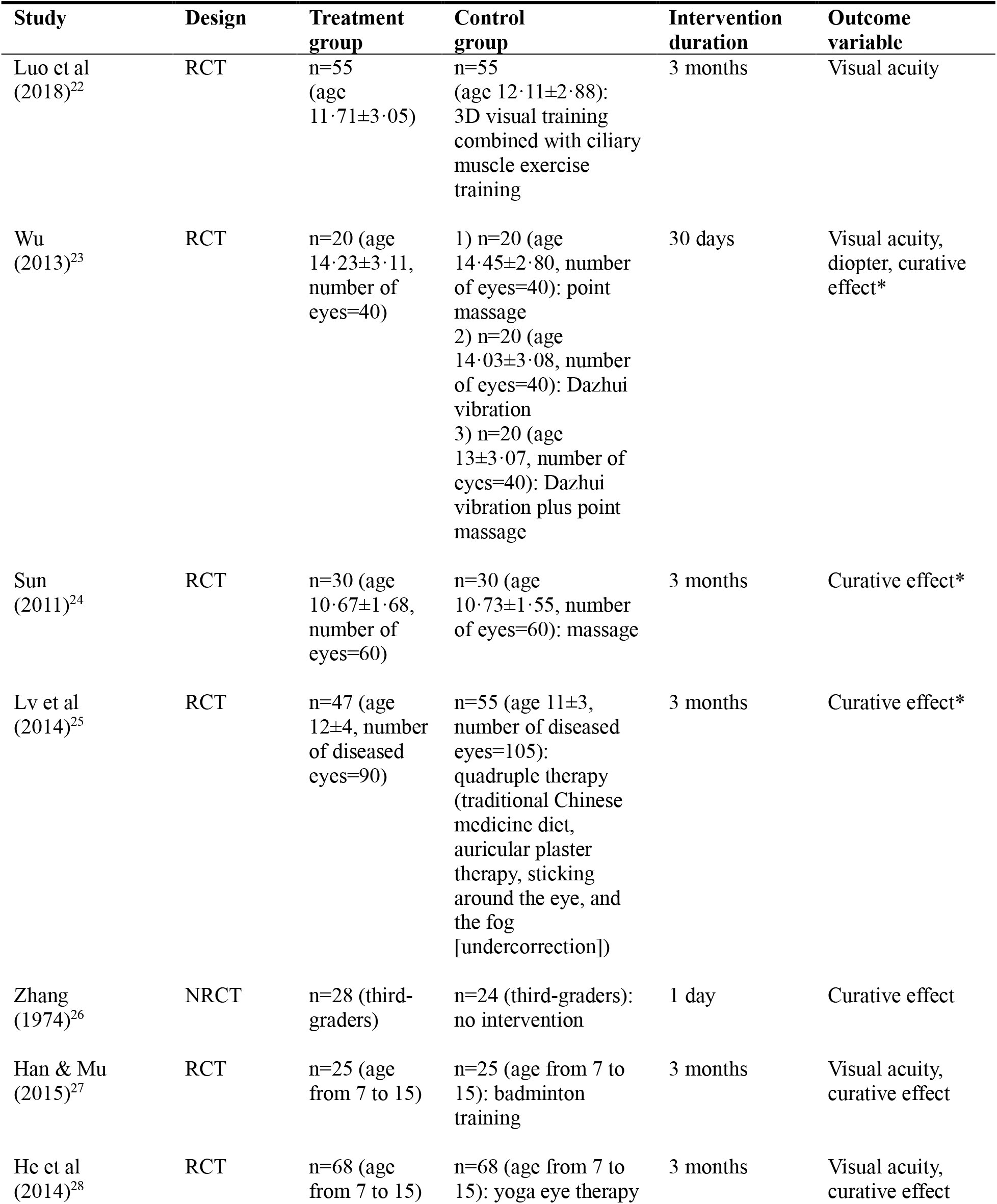
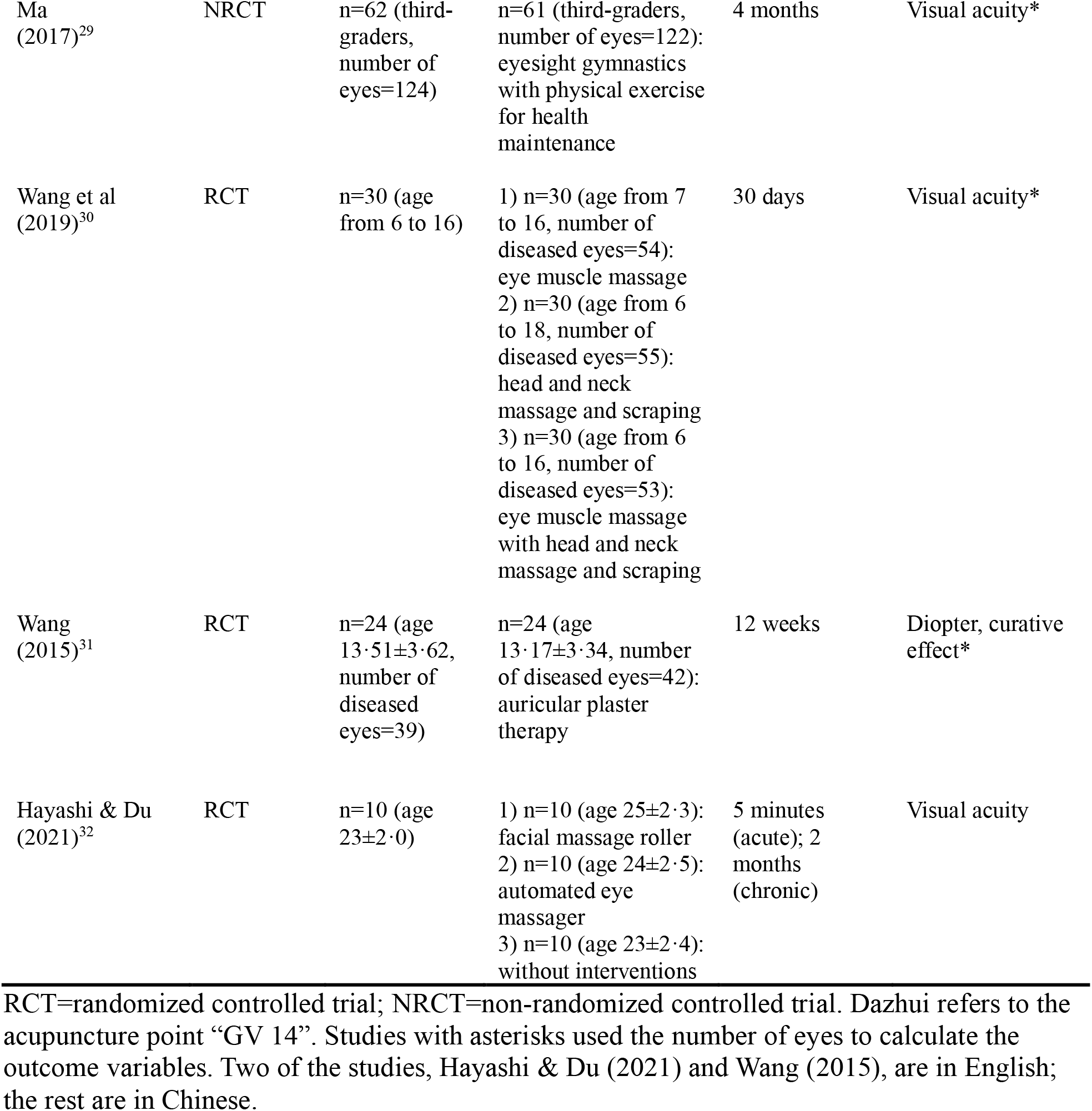
Basic characteristics of the included studies.

**Table: Basic characteristics of the included studies**
| Study | Design | Treatment group | Control group | Intervention duration | Outcome variable |
| --- | --- | --- | --- | --- | --- |
| Luo et al (2018)<sup>22</sup> | RCT | n=55 (age 11·71±3·05) | n=55 (age 12·11±2·88): 3D visual training combined with ciliary muscle exercise training | 3 months | Visual acuity |
| Wu (2013)<sup>23</sup> | RCT | n=20 (age 14·23±3·11, number of eyes=40) | 1) n=20 (age 14·45±2·80, number of eyes=40): point massage<br>2) n=20 (age 14·03±3·08, number of eyes=40): Dazhui vibration<br>3) n=20 (age 13±3·07, number of eyes=40): Dazhui vibration plus point massage | 30 days | Visual acuity, diopter, curative effect* |
| Sun (2011)<sup>24</sup> | RCT | n=30 (age 10·67±1·68, number of eyes=60) | n=30 (age 10·73±1·55, number of eyes=60): massage | 3 months | Curative effect* |
| Lv et al (2014)<sup>25</sup> | RCT | n=47 (age 12±4, number of diseased eyes=90) | n=55 (age 11±3, number of diseased eyes=105): quadruple therapy (traditional Chinese medicine diet, auricular plaster therapy, sticking around the eye, and the fog [undercorrection]) | 3 months | Curative effect* |
| Zhang (1974)<sup>26</sup> | NRCT | n=28 (third-graders) | n=24 (third-graders): no intervention | 1 day | Curative effect |
| Han & Mu (2015)<sup>27</sup> | RCT | n=25 (age from 7 to 15) | n=25 (age from 7 to 15): badminton training | 3 months | Visual acuity, curative effect |
| He et al (2014)<sup>28</sup> | RCT | n=68 (age from 7 to 15) | n=68 (age from 7 to 15): yoga eye therapy | 3 months | Visual acuity, curative effect |
| Ma (2017)<sup>29</sup> | NRCT | n=62 (third-graders, number of eyes=124) | n=61 (third-graders, number of eyes=122): eyesight gymnastics with physical exercise for health maintenance | 4 months | Visual acuity* |
| Wang et al (2019)<sup>30</sup> | RCT | n=30 (age from 6 to 16) | 1) n=30 (age from 7 to 16, number of diseased eyes=54): eye muscle massage<br>2) n=30 (age from 6 to 18, number of diseased eyes=55): head and neck massage and scraping<br>3) n=30 (age from 6 to 16, number of diseased eyes=53): eye muscle massage with head and neck massage and scraping | 30 days | Visual acuity* |
| Wang (2015)<sup>31</sup> | RCT | n=24 (age 13·51±3·62, number of diseased eyes=39) | n=24 (age 13·17±3·34, number of diseased eyes=42): auricular plaster therapy | 12 weeks | Diopter, curative effect* |
| Hayashi & Du (2021)<sup>32</sup> | RCT | n=10 (age 23±2·0) | 1) n=10 (age 25±2·3): facial massage roller<br>2) n=10 (age 24±2·5): automated eye massager<br>3) n=10 (age 23±2·4): without interventions | 5 minutes (acute); 2 months (chronic) | Visual acuity |

RCT=randomized controlled trial; NRCT=non-randomized controlled trial. Dazhui refers to the acupuncture point “GV 14”. Studies with asterisks used the number of eyes to calculate the outcome variables. Two of the studies, Hayashi & Du (2021) and Wang (2015), are in English; the rest are in Chinese.

Overall, out of the 11 studies, nine had some concerns of bias in at least two domains, and two studies had high risk of bias in two domains (appendix p 10). Intercoder consistency in bias rating was high for the first five domains: random sequence generation (% of agreement=100%, PABAK=1, 95% CI 1–1, p<0·001); allocation concealment (% of agreement=95·5%, PABAK=0·90, 95% CI 0·67–1, p<0·001); blinding of participants and personnel (% of agreement=77·3%, PABAK=0·49, 95% CI 0·09–0·88, p=0·02); blinding of outcome assessment (% of agreement=90·9%, PABAK=0·80, 95% CI 0·49–1, p<0·01); incomplete outcome data (% of agreement=86·4%, PABAK=0·69, 95% CI 0·20–1, p=0·01).

Consistency was low for the final two domains: selective reporting (% of agreement=59·1%, PABAK=0·08, 95% CI –0·38 to 0·54, p=0·71) and other biases (% of agreement=54·5%, PABAK=–0·02, 95% CI –0·25 to 0·21, p=0·83). Thus, to better assess the effect of the eye-exercise intervention, sensitivity analysis was subsequently conducted with and without the two high-risk studies.^25,29^

The results from the risk of bias assessments were corroborated by the funnel plots (appendix p 11), which are intended to evaluate the overall publication bias within each outcome measure: visual acuity (within-group and between-group), diopter, and curative effect. Owing to the small number of included studies and the somewhat subjective nature of funnel plots, the results can only be suggestive. Nevertheless, the patterns hint at the presence of publication bias: the distributions are asymmetrical, and the proportion of studies that lie outside the outer lines (where 95% of studies are expected to stay in the absence of biases and heterogeneity) is high, ranging from 22% (curative effect) to 100% (diopter).

Seven studies used visual acuity to measure myopia. Acuity was converted to a common decimal scale. The effect of the eye-exercise intervention was evaluated in one of two ways: 1) within-group changes following intervention (i.e., before and after the eye-exercise intervention); 2) between-group differences following different interventions (i.e., eye-exercise intervention vs. control). For within-group changes (figure 2A), data include 168 participants (218 eyes) from eight experiments. The duration of the intervention ranged from 5 minutes to 4 months. Across experiments, visual acuity declined after the eye-exercise intervention (SMD=–0·67, 95% CI – 1·28 to –0·07, *Z*=2·17, p=0·03). The study heterogeneity was high (*I*^*2*^=93%), and one study^23^ was found to greatly contribute to the high heterogeneity. After excluding this study, the *I*^*2*^ declined to 8%, and the combined effect size became not significant (SMD=–0·12, 95% CI – 0·28 to 0·04, *Z*=1·51, p=0·13). Another study^29^ was found to be of high bias; after excluding it, the combined effect size was significant (SMD=–0·81, 95% CI –1·56 to –0·07, *Z*=2·13, p=0·03). The pattern was the same when both studies were excluded (SMD=–0·24, 95% CI –0·43 to – 0·06, *Z*=2·54, p=0·01). For between-group differences (figure 2A), data include 208 participants (406 eyes) in the eye-exercise groups and 208 participants (404 eyes) in the control groups from 16 different experiments. Before interventions, the two types of groups had comparable visual acuity in each of the experiments. After interventions, across experiments, visual acuity remained similar between the two types of groups (SMD=–0·50, 95% CI –1·16 to 0·16, *Z*=1·49, p=0·14), but was somewhat higher in the eye-exercise groups when the study with adult participants^32^ was excluded (SMD=–1·21, 95% CI –1·98 to –0·43, *Z*=3·04, p<0·01). The heterogeneity was high as well (*I*^*2*^=96%), but no single study contributed to the heterogeneity. One study^29^ was found to be of high bias; after excluding it, the combined effect size remained not significant (SMD=–0·44, 95% CI –1·19 to 0·31, *Z*=1·14, p=0·25).

**Figure 2:**
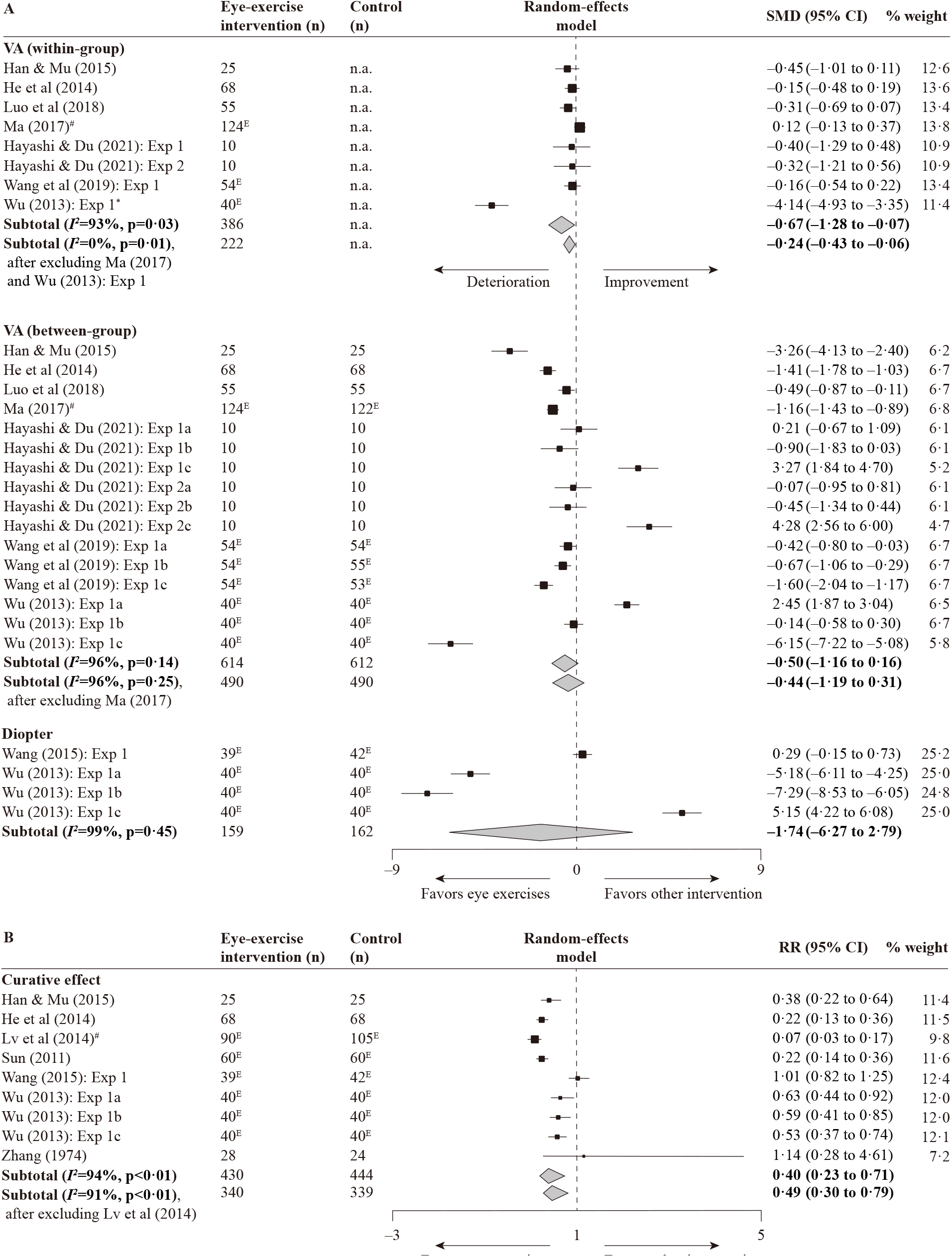
Forest plots for the four comparisons. The dashed line represents a null effect. SMD=standardized mean difference; CI=confidence intervals; n.a.=not applicable; Exp = Experiment; ^E^=eyes (instead of participants); ^*^=study that contributes to high heterogeneity; ^#^=study of high-risk of bias. In the Hayashi & Du (2021) study, Experiments 1 and 2 refer to acute (short-term) and chronic (long-term) experiments, respectively; letters a, b and c refer respectively to comparisons with the no-intervention control, facial massage roller intervention, and automated eye massage intervention. In the Wang et al (2019) study, letters a, b and c refer respectively to comparisons with the eye muscle massage, head and neck massage and scraping, and combined intervention. In the Wu (2013) study, letters a, b and c refer respectively to comparisons with the point massage, Dazhui vibration intervention, and combined intervention.

Two studies used diopter to measure myopia (figure 2A). Data include 159 eyes in the eye-exercise groups and 162 eyes in the control groups from four experiments. Across experiments, the effects of interventions did not differ from each other (SMD=–1·74, 95% CI – 6·27 to 2·79, *Z*=0·75, p=0·45). The study heterogeneity was high (*I*^*2*^=99%), but no single study could be identified to have contributed to it.

Seven studies reported curative effects that evaluated relative risk (figure 2B). Data include 121 participants (309 eyes) in the eye-exercise groups and 117 participants (327 eyes) in the control groups from nine experiments. Across experiments, eye exercises had a higher curative effect than control (RR=0·40, 95% CI 0·23–0·71, *Z*=3·13, p<0·01). Again, the study heterogeneity was high (*I*^*2*^=94%), but no single study could be identified to have contributed to it. One study^25^ was found to be of high bias; after excluding it, the combined effect size was reduced but remained significant (RR=0·49, 95% CI 0·30–0·79, *Z*=2·94, p<0·01).

## Discussion

Eye-exercise interventions were not effective in preventing or controlling the progression of myopia, as indicated by changes in visual acuity and diopter. While there was a reduced risk in eye-exercise groups compared to the control, these studies were highly heterogeneous and might be subject to publication bias. They were also infected with five major weaknesses: a lack of measurement of axial length; small sample sizes (most with n<100 per group); potential biases; failure to consider side effects; and without including established effective interventions as control. The weaknesses of these studies and the lack of empirical evidence do not support the continued use of eye exercises to prevent or control myopia in schoolchildren. Policymakers therefore should abandon this longstanding practice and instead seek evidence-based solutions that are available.

The current meta-analysis examines controlled trials to provide causal evidence on the efficacy of eye exercises in controlling myopia. This builds upon previous indirect and observational evidence, such as the fact that in mainland China, despite the mandatory nationwide practice of eye exercises, the rate of myopia among young adults has steadily increased from 20–30% in the 1980s to 80–90% today, similarly to other East Asian regions and Singapore without the eye-exercise policy.^3^ Indeed, the prevalence of myopia is much higher in China than in many regions without such intervention (eg, Australia). Within China, the myopia rate in urban students, who have much better compliance with the policy, is also much higher compared with rural students^33^—roughly 96.6% of the urban students regularly perform eye exercises,^34^ compared with 15% in rural students.^35^ Other observational studies have also produced little evidence for the effectiveness of eye exercises.^36^ For example, one study found a modest effect on relieving near vision symptoms but no effect on reducing myopia,^34^ a pattern contradicted by a different study;^35^ another study found a statistically significant but probably clinically insignificant effect in reducing accommodative lag;^37^ and still another study found no association between eye exercises and the risk of myopia onset.^38^

In contrast, studies have provided strong support for other interventions to control myopia. Robust evidence indicates that spending time outdoors can protect against developing myopia, in interventions directed at schools^39^ and at families,^40,41^ even though its exact mechanisms of action and its effect on delaying the progression of myopia are not yet settled.^42^ A distinct advantage of outdoor-time intervention is the added benefit of promoting an active lifestyle that helps to enhance mental and physical health more generally. Interventions targeting outdoor time agree with our current understanding of the etiology of myopia: changes in lifestyle over the past several decades, particularly decreased time spent outdoors, likely have played a major role.^3^ Other risk factors, like increased near-work time and education pressure, may also contribute by reducing outdoor time.^3^ Outdoor-time intervention can also be combined with non-lifestyle interventions, such as low-dose atropine and optical interventions, which have been shown to slow myopia progression.^43^

Given the lack of supporting evidence for the continued use of eye exercises and the robust evidence for the effectiveness of interventions such as increased time outdoors, it becomes challenging to justify maintaining the eye-exercise policy. One may argue that students did not perform eye exercises properly—not knowing the correct pressure, the correct location of acupoints, or the basic massage manipulation.^38^ But this misses the larger issue: if after more than 50 years of implementation and most students still cannot do it properly, should we fault the students or the intervention? One may also claim that while eye exercises may not help myopia, they could reduce eye fatigue, or at least pose no harm anyway. This argument overlooks potential inflammation and disease spreading from dirty fingers, as well as the missed opportunity to engage in health-promoting activities such as outdoor play or rest. To justify maintaining the status quote, then, requires robust evidence to show the effectiveness of eye exercises in controlling myopia, ideally from studies that measure axial length, use large sample sizes, minimize biases, examine side effects, and use outdoor time as a control intervention.

Our study has both strengths and limitations. Strengths include the comprehensive evaluation of studies published in both Chinese and English up until December 15, 2022, and the use of controlled trials for more robust causal inference. Limitations include high heterogeneity among studies, the possibility of publication bias, and weaknesses in individual studies such as small sample sizes. Additionally, insufficient studies precluded subgroup analysis for different age groups and for separate evaluations of myopia prevention and control.

In conclusion, eye exercises are not effective in preventing myopia or slowing its progression, as measured by changes in visual acuity and diopter; a small positive effect is observed in curative effects. Given the limited evidence of their effectiveness and the strong evidence supporting alternative interventions such as increased outdoor time, and considering the adverse impacts of COVID-19 lockdown measures in mainland China over the past three years, it is time for policymakers to retire the eye-exercise policy and redirect the time and resources saved toward evidence-based interventions—for the betterment of schoolchildren’s health and the success of myopia control policies.

## Data Availability

Extracted data and code are available online (https://osf.io/dr5jk/); additional requests may be made to the corresponding author.

## Contributors

ZL conceived and designed the study. WC and FX searched the literature and screened articles. FX and WC analyzed the data, constructed the table, and drew the figures under the supervision of ZL. ZL wrote the manuscript. FX and WC accessed and verified the data. FX and WC contributed equally to this work. Yichen Wu and Xinran Ge contributed to the literature search, screening, and coding; Xiani Jia contributed to the editing of a preliminary draft of methods and results. All authors approved the final version of the manuscript and were responsible for the decision to submit the manuscript.

## Acknowledgments

The study was supported by the Guangdong Basic and Applied Basic Research Foundation (2019A1515110574) and Shenzhen Fundamental Research Program (JCYJ20210324134603010). The funding bodies had no role in the study design, data collection, analysis, and interpretation, report writing, or the decision to submit for publication.

## References

1 Holden BA, Fricke TR, Wilson DA, et al. Global prevalence of myopia and high myopia and temporal trends from 2000 through 2050. Am J Ophthalmol 2016; 123(5): 1036–42.

2 Morgan IG, Rose KA. Myopia: is the nature-nurture debate finally over? Clin Exp Optom 2019; 102(1): 3–17.

3 Morgan IG, French AN, Ashby RS, et al. The epidemics of myopia: aetiology and prevention. Prog Retin Eye Res 2018; 62: 134–49.

4 Chua SY, Sabanayagam C, Cheung YB, et al. Age of onset of myopia predicts risk of high myopia in later childhood in myopic Singapore children. Ophthalmic Physiol Opt 2016; 36(4): 388–94.

5 Hu Y, Ding X, Guo X, Chen Y, Zhang J, He M. Association of age at myopia onset with risk of high myopia in adulthood in a 12-year follow-up of a Chinese cohort. JAMA Ophthalmol 2020; 138(11): 1129–34.

6 Ikuno Y. Overview of the complications of high myopia. Retina 2017; 37(12): 2347–51.

7 Naidoo KS, Fricke TR, Frick KD, et al. Potential lost productivity resulting from the global burden of myopia: systematic review, meta-analysis, and modeling. Am J Ophthalmol 2019; 126(3): 338–46.

8 Burton MJ, Ramke J, Marques AP, et al. The Lancet global health commission on global eye health: vision beyond 2020. Lancet Glob Health 2021; 9(4): e489–e551.

9 National health commission of the People’s Republic of China: the 2020 national survey on myopia among children and adolescents. July 13, 2021. http://www.gov.cn/xinwen/2021-07/13/content_5624709.htm (accessed January 21 2023).

10 He M, Huang W, Zheng Y, Huang L, Ellwein LB. Refractive error and visual impairment in school children in rural southern China. Am J Ophthalmol 2007; 114(2): 374–82.

11 Wang H, Li Y, Qiu K, et al. Prevalence of myopia and uncorrected myopia among 721,032 schoolchildren in a city-wide vision screening in southern China: the Shantou myopia study. Br J Ophthalmol 2022.

12 Ma X, Zhou Z, Yi H, et al. Effect of providing free glasses on children’s educational outcomes in China: cluster randomized controlled trial. BMJ 2014; 349: g5740.

13 Wang XQ, Yi HM, Lu LN, et al. Population prevalence of need for spectacles and spectacle ownership among urban migrant children in eastern China. JAMA Ophthalmol 2015; 133(12): 1399–406.

14 Zhu Z, Chen Y, Tan Z, Xiong R, McGuinness MB, Muller A. Interventions recommended for myopia prevention and control among children and adolescents in China: a systematic review. Br J Ophthalmol 2023; 107(2): 160–6.

15 Jan C, Li L, Keay L, Stafford RS, Congdon N, Morgan I. Prevention of myopia, China. Bull World Health Organ 2020; 98(6): 435–7.

16 Rawstron JA, Burley CD, Elder MJ. A systematic review of the applicability and efficacy of eye exercises. J Pediatr Ophthalmol Strabismus 2005; 42(2): 82–8.

17 Morgan IG, Jan CL. China turns to school reform to control the myopia epidemic: a narrative review. Asia Pac J Ophthalmol (Phila) 2022; 11(1): 27–35.

18 Li M, Xu L, Tan CS, et al. Systematic review and meta-analysis on the impact of COVID-19 pandemic-related lifestyle on myopia. Asia Pac J Ophthalmol (Phila) 2022; 11(5): 470–80.

19 Wang J, Li Y, Musch DC, et al. Progression of myopia in school-aged children after COVID-19 home confinement. JAMA Ophthalmol 2021; 139(3): 293–300.

20 Hu Y, Zhao F, Ding X, et al. Rates of myopia development in young Chinese schoolchildren during the outbreak of COVID-19. JAMA Ophthalmol 2021; 139(10): 1115–21.

21 Moher D, Liberati A, Tetzlaff J, Altman DG, Group P. Preferred reporting items for systematic reviews and meta-analyses: the PRISMA statement. PLoS Med 2009; 6(7): e1000097.

22 Luo X, Liu X, Chen Z, Huang X, Chen H. Application research of 3D visual training combined with ciliary muscle exercise training for juvenile myopia rectification. Nursing Practice and Research 2018; 15(03): 90–1.

23 Wu L. Clinical observation on treating juvenile myopia by Dazhui vibrating manipulation plus point massage. 2013.

24 Sun J. The clinical research of applying tonifying spleen Qi Tuina method in treating spleen Qi deficiency type of pediatric myopia. 2011.

25 Lv Y, Dou S, Chen Y, Hong M, Ke, M. Juvenile myopia treatment by Four Combined methods for 105 eyes. Chin Med Mod Distance Edu 2014; 12(06): 10–1.

26 Zhang L. Observation of the influence of eye exercise on visual functions of pupils. Guangxi Med 1974; (04): 37–8.

27 Han B, Mu T. Observation on the effect of badminton training on adolescent pseudomyopia. Chin J Convalescent Med 2015; 24(02): 174–5.

28 He J, Hao Y, Peng Q, Chen J. Efficacy of yoga eye exercise on juvenile pseudomyopia. J Beihua Univ (Natural Sci Edit) 2014; 15(01): 85–7.

29 Ma Z. Research on the creation and compilation of gymnastic exercise for sports health care and prevention and treatment of myopia in teenagers. 2017.

30 Wang Y, Bao X, Shi Y, Wang Z. Clinical study of muscle massage around eyes combined with scraping of head and neck in the treatment of adolescent pseudomyopia. J Heibei Trad Chin Med and Pharmacol 2019; 34(06): 35–8.

31 Wang YQ. Effect of eye exercises in combination with auricular plaster therapy on adolescent pseudomyopia patients. 3rd AASRI Conference on CIB Korea: 2015.

32 Hayashi N, Du L. Acute and chronic periocular massage for ocular blood flow and vision: a randomized controlled trial. Int J Ther Massage Bodywork 2021; 14(2): 5–13.

33 He MG, Zheng YF, Xiang F. Prevalence of myopia in urban and rural children in Mainland China. Optom Vis Sci 2009; 86(1): 40–4.

34 Lin Z, Vasudevan B, Jhanji V, et al. Eye exercises of acupoints: their impact on refractive error and visual symptoms in Chinese urban children. BMC Complement Altern Med 2013; 13: 306.

35 Lin Z, Vasudevan B, Fang SJ, et al. Eye exercises of acupoints: their impact on myopia and visual symptoms in Chinese rural children. BMC Complement Altern Med 2016; 16: 349.

36 Lu ZP, Ouyang MZ, Zhang R, Tang X, Zhong HJ. Association between Chinese eye exercises and onset of myopia: a meta-analysis. Int J Clin and Exp Med 2019; 12(5): 4580–8.

37 Li SM, Kang MT, Peng XX, et al. Efficacy of Chinese eye exercises on reducing accommodative lag in school-aged children: a randomized controlled trial. Plos One 2015; 10(3): e0117552.

38 Kang MT, Li SM, Peng X, et al. Chinese Eye exercises and myopia development in school age children: a nested case-control study. Sci Rep 2016; 6: 28531.

39 He M, Xiang F, Zeng Y, et al. Effect of time spent outdoors at school on the development of myopia among children in China: a randomized clinical trial. JAMA 2015; 314(11): 1142–8.

40 Li SM, Ran AR, Kang MT, et al. Effect of text messaging parents of school-aged children on outdoor time to control myopia: a randomized clinical trial. JAMA Pediatr 2022; 176(11): 1077–83.

41 Li Q, Guo L, Zhang J, et al. Effect of school-based family health education via social media on children’s myopia and parents’ awareness: a randomized clinical trial. JAMA Ophthalmol 2021; 139(11): 1165–72.

42 Lingham G, Mackey DA, Lucas R, Yazar S. How does spending time outdoors protect against myopia? A review. Br J Ophthalmol 2020; 104(5): 593–9.

43 Wildsoet CF, Chia A, Cho P, et al. IMI - interventions myopia institute: interventions for controlling myopia onset and progression report. Invest Ophthalmol Vis Sci 2019; 60(3): M106–M31.

